# Emergence and co-circulation of three multidrug resistant *Shigella sonnei* strains among men who have sex with men, Portland, Oregon, 2017–2019

**DOI:** 10.1101/2025.10.28.25338796

**Authors:** Russell S. Barlow, Sara McCall, Kimberly A. Schwindt, Chris Nytko, Lisa Ferguson

## Abstract

*Shigella* is a highly infectious bacterial pathogen spread via the fecal-oral route. Person to person spread during sexual contact among men who have sex with men (MSM) has been widely reported (*1, 2*). From January 2017 through May 2019, 74 culture confirmed multidrug resistant (MDR) shigellosis infections were reported in the Portland, Oregon area. Infections were caused by three divergent *Shigella sonnei* strains. To prevent additional illness and characterize the epidemiology of MDR shigellosis, a multifaceted investigation was conducted including expanded questionnaires, whole genome sequencing (WGS), antimicrobial susceptibility testing (AST), medical chart review, and sex partner notification. Among the 68 cases for which interviews were completed, 66 (97%) were among MSM. Patients reported having multiple sex partners (90%) and meeting on mobile dating or hook-up applications (63%). Forty percent reported methamphetamine use. Prevention and control challenges included low awareness of shigellosis among MSM and healthcare providers, inappropriate antimicrobial therapy, treatment failures, and limited sex partner elicitation. Results highlight the need for interventions to increase MSM and provider awareness of MDR *Shigella* spread through sexual contact to reduce shigellosis morbidity, limit ongoing transmission, and prevent future outbreaks.

**Summary:** *What is already known about this topic?:* Multidrug resistant (MDR) shigellosis among men who have sex with men (MSM) has been widely reported. Risk factors for specific MDR strains are of concern.

*What is added by this report?:* Between 2017 and 2019, three diverse MDR *Shigella sonnei* strains began co-circulating among sexually active MSM in the Portland, Oregon area. Patients reported multiple sex partners, use of mobile applications to meet partners, methamphetamine use, and housing insecurity. MDR shigellosis commonly resulted in hospitalization, treatment failure, and prolonged morbidity.

*What are the implications for public health practice?:* Interventions to increase MSM and provider awareness of MDR *Shigella* spread through sexual contact are necessary. To reduce morbidity and limit transmission, sexual health education should be provided to all adult patients with *Shigella*, and healthcare providers should order culture with antimicrobial susceptibility testing to inform antimicrobial therapy.

## Investigation and Outcomes

During August 2017, the rate of domestically acquired shigellosis increased in the Portland, Oregon area. Primary interviews indicated that all cases were in males that endorsed sexual contact with another male in the week prior to illness onset. AST indicated multi-drug resistance including reduced susceptibility to ciprofloxacin. To prevent additional illness and characterize the epidemiology of MDR shigellosis, Portland, Oregon area public health partners implemented expanded *Shigella* questionnaires, 2) WGS, 3) medical chart review, 4) AST, and 5) sex partner notification. Confirmed MDR Shigellosis was defined as isolation of *Shigella sonnei* with demonstrated or predicted *(3)* resistance to at least two antibiotic classes, including reduced susceptibility to fluoroquinolones (ciprofloxacin minimum inhibitory concentration [MIC] ≥0.50 ug/ml) or 3^rd^ generation cephalosporins (ceftriaxone MIC ≥4 ug/ml) (*4, 5*). Expanded questionnaires asked about a range of clinical, demographic, and behavioral factors. We defined MSM as any adult male who self-identified as homosexual or bisexual or that reported sexual contact with another male in the three months prior to illness onset. Demographic and behavioral characteristics of patients were compared across strains using the Fisher’s exact test or the Kruskal-Wallis test; *P* values ≤ 0.05 were considered statistically significant.

From January 2017 through May 2019, 74 sporadic, confirmed infections with MDR *Shigella sonnei* were reported in the Portland, Oregon area including Multnomah, Clackamas, and Washington Counties. Epidemiological and WGS data indicated that these infections comprised five separate clusters among three divergent MDR strains (Figure 1). Strain 1 (n=42) exhibited resistance to all 1^st^ line antibiotics except ceftriaxone (Table 1). WGS and epidemiological data indicated that this strain was introduced at least five separate times and resulted in three propagated clusters. The first introduction occurred in 2017 (n=5), followed by a second introduction in May 2018 of a related strain that was 22-31 single nucleotide polymorphisms (SNP) apart (n=17). During August 2018, a related strain (7-50 SNPs apart from the other clusters) was identified (n=18) and this introduction ultimately resulted in a restaurant associated foodborne outbreak in December 2018 (n=6 additional cases, not included in this report). Strain 2 (n=14) was susceptible to ampicillin and ceftriaxone and represented a single cluster. Finally, Strain 3 (n=18) caused a single cluster and was an extended-spectrum beta-lactamase producer (ESBL, *bla*CTX-M-27).

**Table 1.** Genetic and antimicrobial susceptibility profiles of multidrug resistant sporadic *Shigella sonnei* isolates from the Portland Oregon area, January 2017 through May 2019 (N=74).

|  | Strain 1 | Strain 2 | Strain 3 |
| --- | --- | --- | --- |
| First Reported | February, 2017 | February, 2018 | November, 2018 |
| Total number of isolates | 42 | 14 | 18 |
| Number of Clusters | 3 | 1 | 1 |
| SNP Range* | 0-50 | 0-2 | 0-11 |
| Number (%) with AST performed | 34 (81%) | 12 (86%) | 15 (83%) |
| **Antibiotics†** | **Resistant** | **Resistant** | **Resistant** |
| Ampicillin | 100 | 0 | 100 |
| Azithromycin (% with *erm(B)* and *mph(A)*)§ | 100 | 100 | 100 |
| Ceftriaxone | 0 | 0 | 100 |
| Ciprofloxacin¶ (Median Minimum Inhibitory Concentration [MIC] Value) | 100 (>1ug/ml) | 100 (0.50ug/ml) | 0 (0.12ug/ml) |
| Sulfamethoxazole-trimethoprim | 100 | 100 | 100 |
| Selected Resistance Mechanisms Detected (3) | *gyrA* 83, *gyrA* 87, *parC* 80 mutations mediating fluoroquinolone resistance, *erm(B)* and *mph(A)* | *qnrS13* mediated fluoroquinolone resistance, *erm(B)* and *mph(A)* | Extended spectrum beta lactamase producer (ESBL, *bla*<sub>CTX-M-27</sub>), *erm(B)* and *mph(A)*, gyrase A mutation (*gyrA*-S83L) |

\*Single nucleotide polymorphism (SNP)
† CLSI Breakpoints were used for ampicillin, ceftriaxone, and sulfamethoxazole-trimethoprim (4)
§Inferred by the presence of both macrolide resistance factors *erm(B)* and *mph(A)*, as determined from The NCBI Pathogen Detection Project (2, 3)
¶MIC of ≥0.50ug/ml used as the breakpoint for fluoroquinolone reduced susceptibility (4, 5)

**Figure 1.**
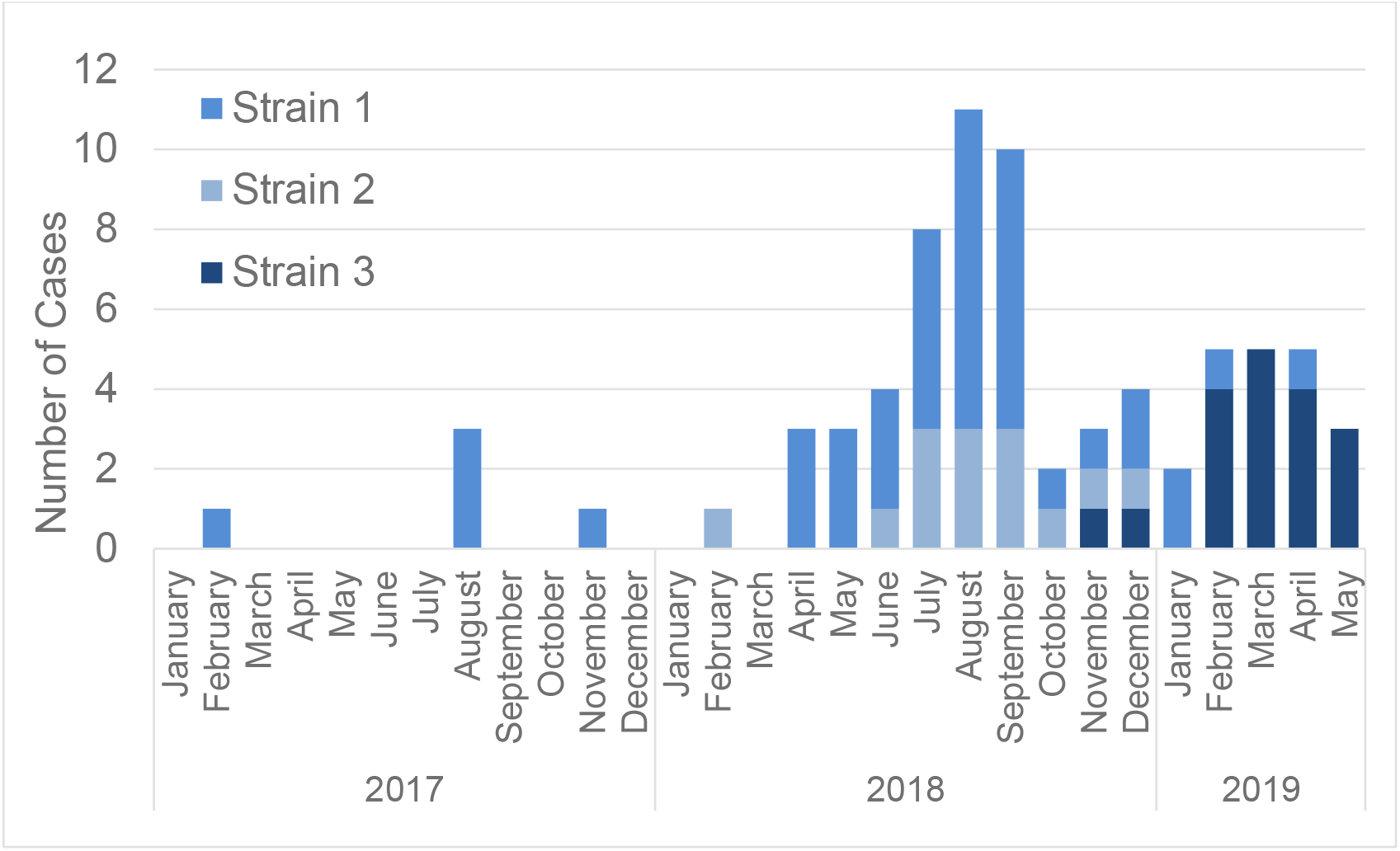
Multidrug resistant *Shigella sonnei* reported illness onset by month. Portland, Oregon area, January 2017 through May 2019 (N=74).

Interviews were completed for sixty-eight (92%) cases. None reported international travel and no point or common source exposures were identified. Age, sex, and race/ethnicity distributions were comparable across strains. Median age was 42.5 years (interquartile range [IQR] 34-53 years). Sixty-seven (99%) patients were male; 56 (82%) identified as white, non-Hispanic, 4 (6%) were white, Hispanic, 3 (4%) were Asian, 1 (1%) was Black, and 4 (6%) were of mixed racial identities. Sixty-six (97%) patients were MSM and 32 (47%) were HIV seropositive (Table 2). Qualitatively, case investigators noted that patients had low awareness of the sexual spread of shigellosis.

**Table 2.**
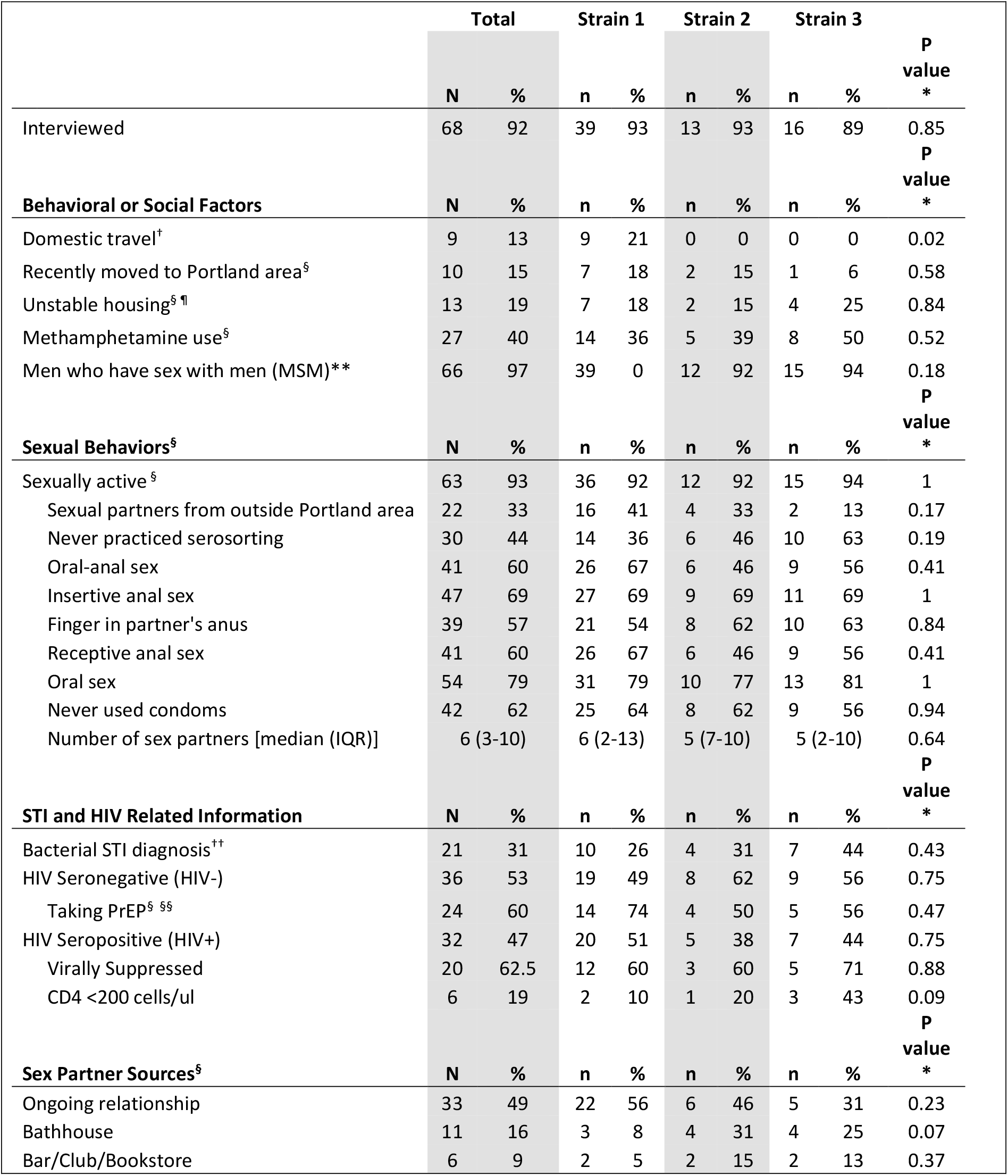
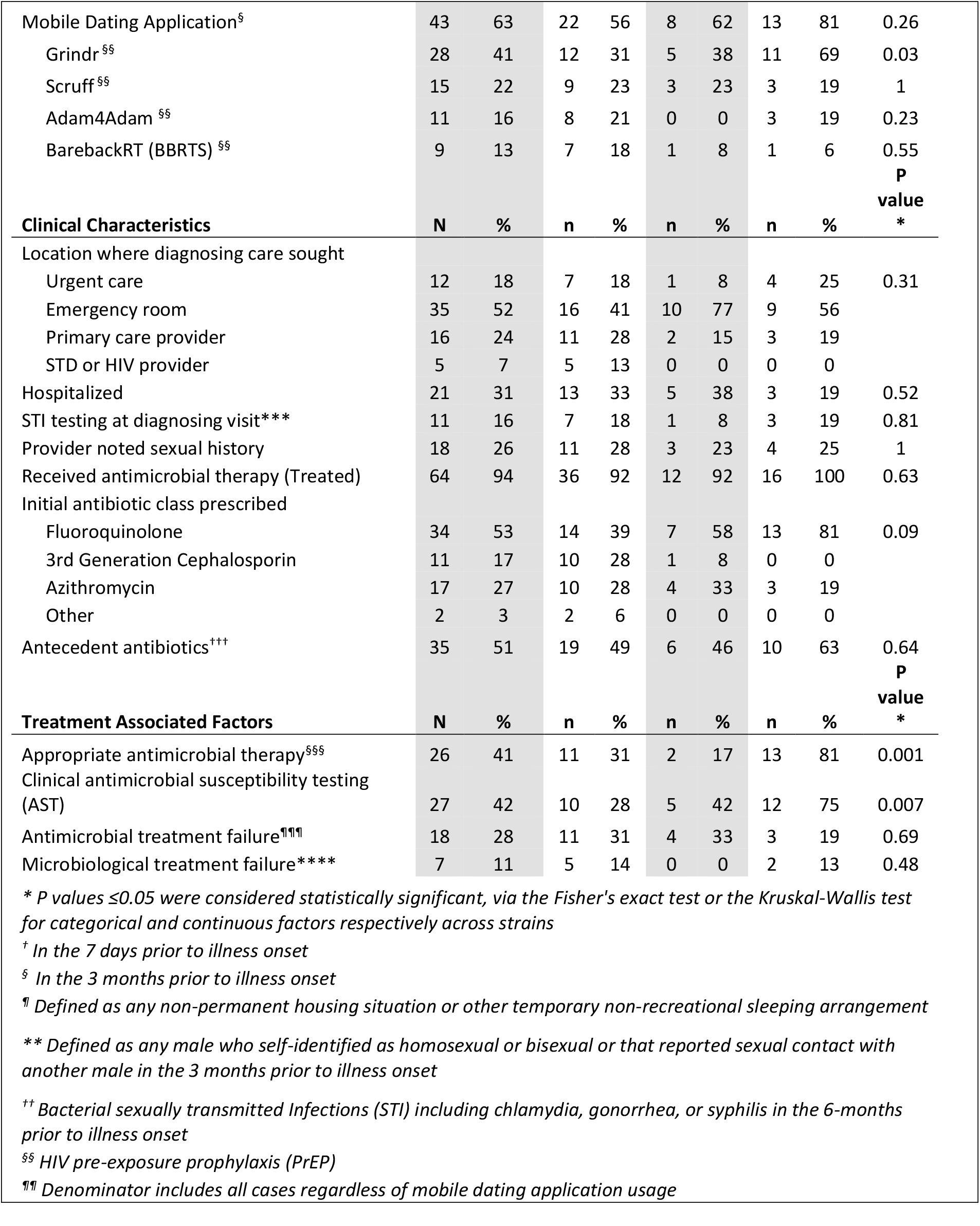
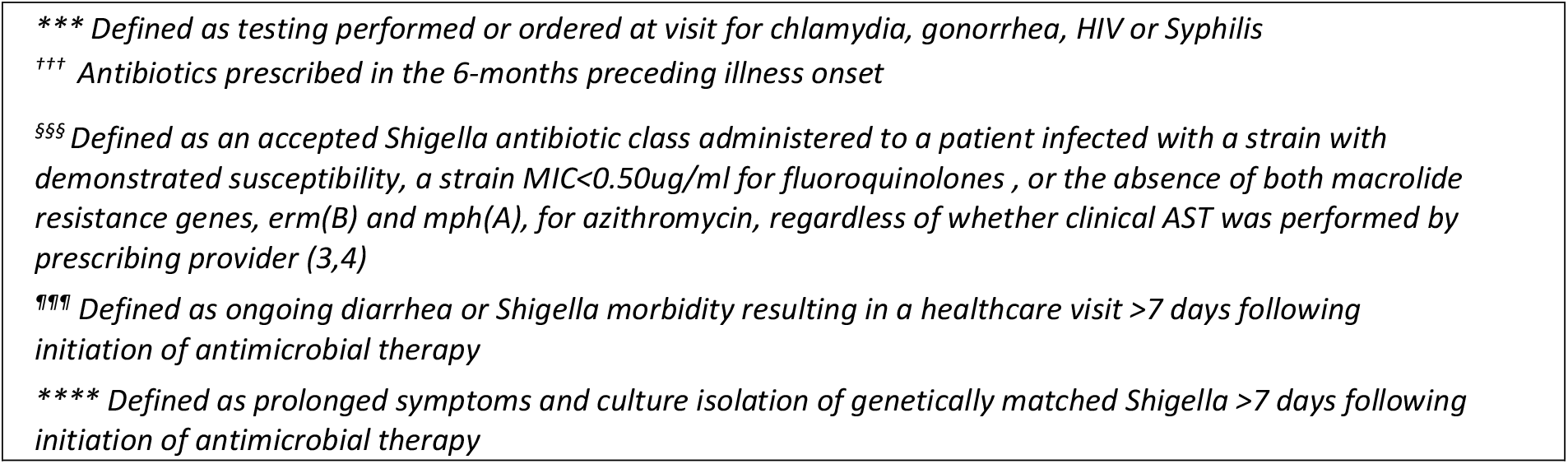
Behavioral, social, and clinical characteristics of patients with multidrug resistant *Shigella sonnei* isolates, Portland Oregon area, January, 2017 through May, 2019 (N=68).

Behavioral characteristic distributions were similar across strains. However, strain-specific social mixing patterns were identified including mobile applications, domestic travel, and possibly bathhouse attendance (Table 2). Relevant overlapping characteristics in the three months prior to illness onset included multiple sex partners (median 6, IQR 3-10) often facilitated through use of mobile applications (n=43, 63%). Forty-one (60%) patients reported oral-anal sexual contact, 42 (62%) denied using condoms, 30 (44%) never practiced serosorting, 27 (40%) used methamphetamine including sexualized drug use (e.g., chem-sex), and 21 (31%) had recent bacterial sexually transmitted infection (STI) diagnoses. Partner elicitation was low overall, with only 18 (26%) patients naming partners. Sexual partner notification identified additional cases (n=6). No patients named partners that had already been identified through disease reporting.

Interviews and medical chart review indicated that cases were primarily diagnosed in acute care settings (Table 2). According to case investigators, patients reported that providers rarely discussed sexual risk or prevention of shigellosis. Indeed, provider-documented patient sexual histories and STI testing were uncommon. Clinical outcomes were similar across strains.

Overall, 21 (31%) patients were hospitalized and 64 (94%) received antimicrobial therapy. AST and appropriate therapy differed by strain. Collectively, only 41% (n=26) of treated patients had clinical AST performed on their isolate, with 81% (n=21) of AST performed by a single health system. Lack of AST and clinical breakpoints for azithromycin resulted in at least 38 (58%) patients receiving antibiotics that were likely inappropriate. Consequently, 18 (29%) treated patients experienced prolonged morbidity or clinical treatment failure^*^, including 14 individuals that received inappropriate antimicrobial therapy (odds ratio for treatment failure for those with inappropriate therapy 3.2, 95% confidence interval 0.9-11.2, p value 0.09). Patients with clinical treatment failures utilized additional healthcare (24 visits), and four patients with treatment failures were subsequently hospitalized. All individuals with clinical treatment failure^†^ that were tested remained culture positive (n=7, 39% of all treatment failures). Positivity persisted for a minimum median^~^ of 49 days from illness onset (range 17-96 days).

## Discussion

During 2017-2019, the Portland, Oregon area experienced the emergence and co-circulation of multiple, highly drug resistant *Shigella sonnei* strains disproportionately affecting MSM. The increased dissemination of MDR *Shigella* was likely being driven by behavioral factors identified here and reflect findings reported elsewhere (*1, 6, 7, 8*). These include multiple sex partners facilitated by mobile dating apps and sex venues, sex partners from outside the Portland area, lack of *Shigella* awareness among MSM, methamphetamine use including sexualized drug use, and routine exposure to clinically relevant antibiotics.

Poor health outcomes and prolonged shedding may have been exacerbated by healthcare-associated contributory factors including inadequate provider awareness of the spread of *Shigella* and the high level of antimicrobial resistance. Providers infrequently noted sexual history or tested for other STIs at diagnosing visits. Therefore, lack of sexual health education and inappropriate antibiotic use may have contributed to *Shigella* transmission among MSM who engage in sexual behavior and are unaware of the ongoing risk to partners.

This investigation had multiple limitations and challenges. Specifically, behavioral data help define the populations impacted but do not necessarily correspond to risk of *Shigella* acquisition. Similarly, sex partner elicitation rates were low, in part, due to app and venue facilitated anonymous sex, and were very resource intensive. As a result, community engagement including gay and bisexual men key informant interviews were performed to define effective outreach and prevention messaging. This study may underestimate total disease burden and overestimate severity, since reported cases likely represent only those with most severe illness. Therefore, predictors of severe and prolonged *Shigella* infections are being analyzed to inform future clinical intervention strategies. Relatedly, provider outreach to increase AST proved ineffective since most health systems no longer support routine enteric AST. Consequently, case investigators routinely caution about antibiotic resistance and recommend follow-up testing to empower behavior modification and confirm *Shigella* clearance.

The increasing population prevalence of shigellosis among MSM combined with intersecting related factors (e.g., illicit drug use, housing insecurity, travel, work in healthcare, daycare, or food industry) provide opportunities for MDR *Shigella* dissemination. Indeed, strains 1 and 3 caused shigellosis outbreaks at a restaurant in Oregon and a long-term care facility in Vermont, respectively (*9*). Similarly, a previous MSM-associated strain caused an outbreak in the Portland, Oregon area among individuals experiencing homelessness (*10*). Since the writing of this manuscript, strain 3 caused large outbreaks at a restaurant and among persons experiencing homelessness in Portland, Oregon. Therefore, disrupting transmission can reduce morbidity among MSM and prevent future outbreaks among other risk populations.

To limit dissemination of MDR shigellosis and reduce morbidity, healthcare providers should suspect and test for *Shigella* among adult male patients with diarrhea. Diagnosing providers should perform AST for patients prior to administration of antimicrobials, routinely offer follow-up testing, and discuss *Shigella* sexual and behavioral health education after diagnosis. To define the relationship between behavioral factors and MDR *Shigella* dissemination, health departments should routinely collect relevant behavioral and demographic information, provide *Shigella* sexual health education to all Shigellosis case-patient reports and contacts, and engage in population-specific (provider, MSM, etc) *Shigella* outreach and prevention messaging.

## Data Availability

These data are protected by public health law and are not publicly available

## Acknowledgments

The authors would like to acknowledge the contributions of the communicable disease teams at Washington and Clackamas County Health Departments in Oregon, Tyler Boyet at Multnomah County Health Department, the Oregon State Public Health Laboratory Microbiology section, Emilio DeBess at the Oregon Health Authority, and the Portland, Oregon area residents that participate in communicable disease interviews. Additionally, the authors would like to acknowledge Amanda Garcia-Williams PhD, Zachary A. Marsh, MPH, Division of Foodborne, Waterborne, and Environmental Diseases, National Center for Emerging Zoonotic and Infectious Diseases, CDC for their helpful insights, comments, and edits.

## Footnotes

* ***Treatment failure*** *(clinical) is defined here as ongoing diarrhea or Shigella morbidity resulting in a healthcare visit >7 days following initiation of antimicrobial therapy*

† ***Microbiologic treatment failure*** *is defined here as prolonged symptoms and culture isolation of genetically matched Shigella >7 days following initiation of antimicrobial therapy*

† ***Minimum median shedding*** *was the interval from reported symptom onset and the date that the last positive specimen was collected*.

